# Inferring Transmission Fitness Advantage of SARS-CoV-2 Variants of Concern in Wastewater Using Digital PCR

**DOI:** 10.1101/2021.08.22.21262024

**Authors:** Lea Caduff, David Dreifuss, Tobias Schindler, Alexander J. Devaux, Pravin Ganesanandamoorthy, Anina Kull, Elyse Stachler, Xavier Fernandez-Cassi, Niko Beerenwinkel, Tamar Kohn, Christoph Ort, Timothy R. Julian

## Abstract

Throughout the global COVID-19 pandemic, SARS-CoV-2 genetic variants of concern (VOCs) have repeatedly and independently arisen. VOCs are characterized by increased transmissibility, increased virulence, or reduced neutralization by antibodies obtained from prior infection or vaccination. Tracking the introduction and transmission of VOCs relies on sequencing, typically whole-genome sequencing of clinical samples. Wastewater surveillance is increasingly used to track the introduction and spread of SARS-CoV-2 variants through sequencing approaches. Here, we adapt and apply a rapid, high-throughput method for detection and quantification of the frequency of two deletions characteristic of the B.1.1.7, B.1.351, and P.1 VOCs in wastewater. We further develop a statistical approach to analyze temporal dynamics in drop-off RT-dPCR assay data to quantify transmission fitness advantage, providing data similar to that obtained from clinical samples. Digital PCR assays targeting signature mutations in wastewater offer near real-time monitoring of SARS-CoV-2 VOCs and potentially earlier detection and inference on transmission fitness advantage than clinical sequencing.

## INTRODUCTION

Emergence of SARS-CoV-2 genetic variants of concern (VOCs) threaten progress toward reductions in the global disease burden attributable to COVID-19. VOCs emerge when novel mutations in the genome increase transmissibility and/or virulence, or reduce neutralization by antibodies obtained from prior infection or vaccination. Notable examples include Alpha (lineage B.1.1.7, first described in the United Kingdom in September 2020), Beta (lineage B.1.351 first discovered in South Africa in December 2020), and Gamma (lineage P.1 first discovered in Brazil in January 2021). VOCs are identified by combinations of co-occurring mutations. Tracking the introduction and transmission of VOCs relies on sequencing, typically whole genome-sequencing of clinical samples. Given the need for lower cost, easy-to-interpret, higher throughput, and more rapid methods, PCR-based methods targeting signature or defining mutations (typically point mutations or deletions) are increasingly common (Vogels et al. 2021; Bedotto et al. 2021).

Because SARS-CoV-2 RNA is shed in feces and other bodily fluids (Wang et al. 2020; D. L. Jones et al. 2020), it is readily detectable in wastewater samples. Surveillance for SARS-CoV-2 in wastewater (wastewater-based epidemiology) is used to inform COVID-19 disease trajectories within catchment areas. Wastewater can also be used to inform the introduction and spread of SARS-CoV-2 variants. To date, sequencing SARS-CoV-2 in wastewater has been applied to detect novel variants in San Francisco Bay Area (Crits-Christoph et al. 2021), detect and track geographic spread of B.1.1.7 in Switzerland (Jahn et al. 2021), and track rises in frequency of B.1.1.7 in London (Wilton et al. 2021). However, in contrast to clinical samples, wastewater samples consist of a mix of SARS-CoV-2 variants representative of the variants circulating within the catchment areas. Therefore, VOCs in wastewater are inferred by tracking combinations of multiple signature or characteristic mutations. Confidence in detection can be increased by investigating temporal trends and/or tracking signature mutations that co-occur on a single amplicon (Jahn et al. 2021).

Although sequencing SARS-CoV-2 RNA in wastewater is a promising approach to track VOCs from hundred to thousand individuals in a single sample, there are temporal lags in sample collection and analysis, similar to lags associated with clinical testing. Sequencing and bioinformatics pipelines typically take at least 3 days to yield results after sample RNA is collected. For more rapid identification of VOCs, PCR-based assays are developed to identify their characteristic mutations (Vogels et al. 2021; Bedotto et al. 2021; Chaintoutis et al. 2021). Examples include assays targeting single-nucleotide polymorphisms, such as N501Y and E484K in the spike protein, or characteristic deletions such as spike Δ69-70 and ORF1a Δ3675-3677. The PCR-based assays, designed for detection of variants in clinical samples, have also been applied successfully for detection in wastewater (Wurtzer et al. 2021; Lee et al. 2021; Heijnen et al. 2021). Using quantitative PCR-based assays (RT-qPCR or RT-dPCR), the proportion of a specific variant within the population can be inferred. Proportions are determined using quantitative assays by comparing the quantity of the target gene region characteristic of the variant to the quantity of a conserved region either on a distinct gene (i.e., N gene) or on a region adjacent to the VOC-specific region (Wurtzer et al. 2021; Lee et al. 2021; Heijnen et al. 2021). Although this approach has demonstrated correlation with proportions of variants in clinical samples, bias is introduced by variation in efficiency and sensitivity of the quantification approaches of two different assays.

Here, we adapt and apply a rapid, high-throughput method for detection and quantification of the frequency of two deletions characteristic of the B.1.1.7, B.1.351, and P.1 VOCs in wastewater. We quantify proportions of the variant within a wastewater sample using two non-competitive hydrolysis probes targeting conserved regions on the same amplicon within a single RT-dPCR assay (“drop-off RT-dPCR assay”). This allows for simultaneous quantification of wild type (strain not carrying the mutation of interest) and variants. The assay is applied to raw influent obtained from a wastewater treatment plant in Zurich, Switzerland, over the period December 2020 - March 2021 when the B.1.1.7 lineage was introduced and spread throughout the region (Chen, Nadeau, Topolsky, Manceau, Huisman, Jablonski, et al. 2021). We further develop a statistical approach to analyze temporal dynamics in drop-off RT-dPCR assay data to quantify transmission fitness advantage, providing data similar to that obtained from clinical samples. Digital PCR assays targeting signature mutations in wastewater offer near real-time monitoring of SARS-CoV-2 VOCs, and potentially earlier inference on epidemiology of VOC transmission.

## METHODS

### Drop-Off RT-dPCR Assays for Variant of Concern Detection in Wastewater

A first hydrolysis probe-based drop-off RT-dPCR assay was developed to target spike Δ69-70 characteristic of Alpha (lineage B.1.1.7), and a second assay was developed for ORF1a Δ3675-3677 characteristic of Alpha (lineage B.1.1.7), Beta (lineage B.1.351), and Gamma (lineage P.1). Although these deletions are characteristic of VOCs, both have been detected in other variants, including non-VOCs (Vogels et al. 2021). Notably, spike Δ69-70 is associated with increased infectivity (Harvey et al. 2021; Kemp et al. 2021). The assay adapts the approach of Vogels et al. (Vogels et al. 2021) in which the hydrolysis probes anneal only on amplicons without the deletion, and are hereafter referred to as “deletion probes”. Vogels et al. (2021) report specificity of the hydrolysis probes based on RT-qPCR analysis of a suite of 71 previously sequenced clinical samples (Vogels et al. 2021).

To measure the fraction of VOCs, we adapted the assay by designing additional hydrolysis probes, which target conserved regions of the same amplicons that detect the deletion (Table 1). These probes, hereafter referred to as “universal probes”, detect amplicons of all lineages, including those with deletions. Based on the ratio of dPCR droplets with single fluorescence (corresponding to amplicons with a deletion) to double fluorescence (amplicons without the deletion), the proportion of VOCs in the wastewater can be estimated (Figure 1). Further, Alpha variants can be distinguished from Beta and Gamma by comparing dPCR results of the two assays targeting the spike Δ69-70 and ORF1a Δ3675-3677 (Vogels et al. 2021).

**Table 1.**
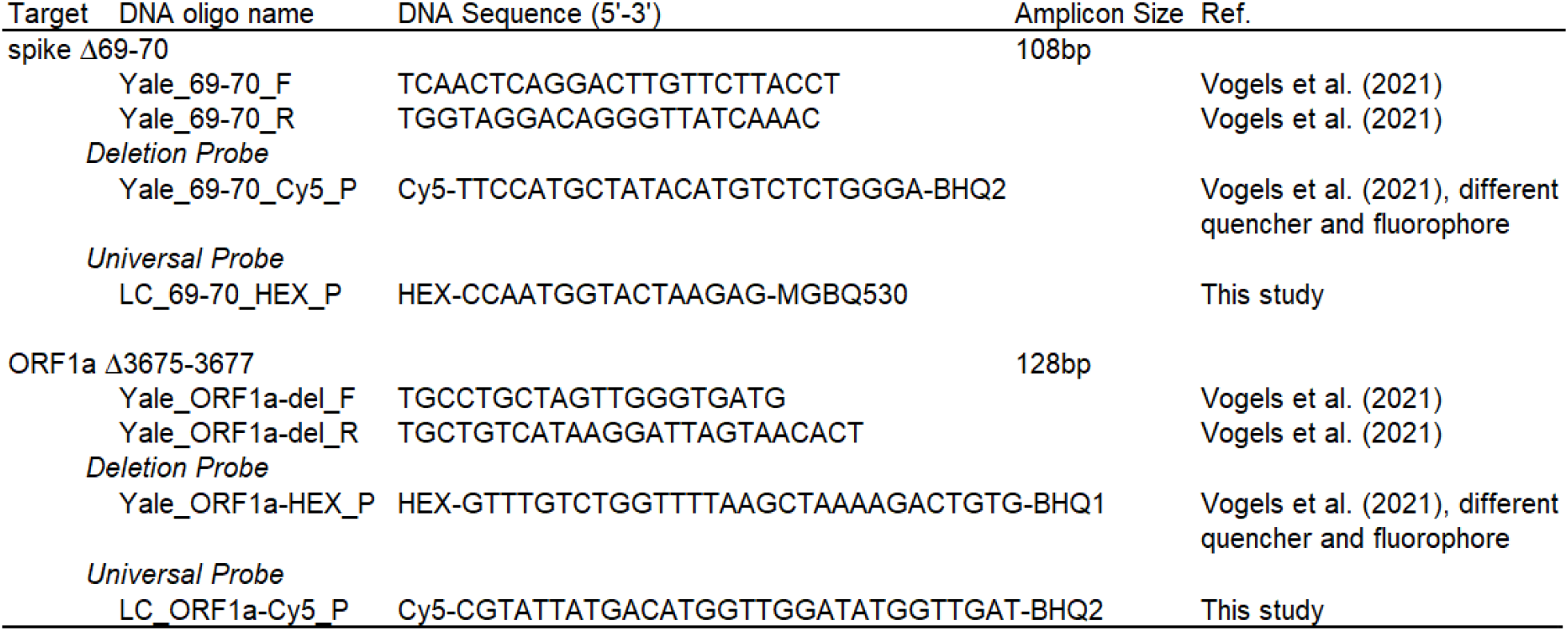
Drop-off RT dPCR assay primer and probe sequences, including resulting amplicon size and corresponding reference.

**Figure 1.**
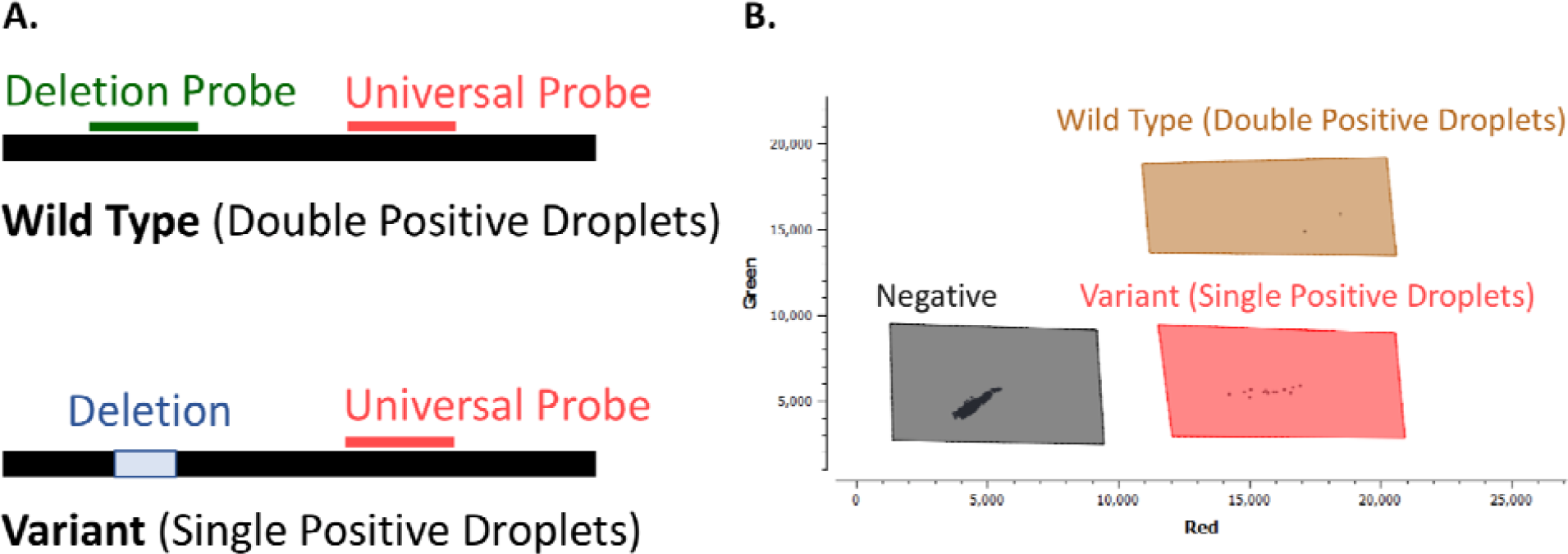
Schematic overview of the drop-off RT-dPCR assay based on two different probes; a deletion probe and a universal probe. A) The universal probe binds to both the wild type and the variant, while the deletion probe only binds to the wild type (no deletion). B) The wild type appears as double positive droplets (brown) and the variant as single positive droplets (red) in the associated dPCR 2D scatterplot.

The drop-off RT-dPCR assays were performed with the Crystal Digital PCR using the Naica System (Stilla Technologies) with the qScript XLT 1-Step RT-PCR Kit (QuantaBio). Reactions were prepared as 27 μl (using 5.4 μl template and 21.6 μl mastermix) pre-reactions, of which 25 μl was loaded into Sapphire Chips (Stilla Technologies) for an equivalent of 5 μl template per reaction. Mastermix composition was slightly different depending on the assay (see Supplemental Material, Table 1). Templates (RNA extracts from wastewater) were diluted 1:2 to maximize the number of gene copies per reaction, which is influenced by sample inhibition (see Supplemental Material, Figure S1). Any measurement was performed in duplicates. Thermocycler conditions included partitioning droplets (40°C for 12 min), reverse transcription (55°C for 30 min), and polymerase activation (95°C for 1 min) followed by 40 cycles of denaturation (95°C for 10 sec) and annealing/extension (55°C for 30 sec).

### Benchmarking Drop-Off RT-dPCR Assay

The drop-off RT-dPCR assay performance was first tested against RNA extracts of the SARS-CoV-2 Wuhan-Hu-1 lineage (representing the wild type sample) and of the B.1.1.7 lineage (representing the VOC sample). RNA extracts from both samples were diluted in molecular grade water to a target concentration of 200 gc/μl. Each hydrolysis probe was first tested in singleplex assays. Then the universal and the deletion probe for each amplicon were combined to yield two duplex assays targeting amplicons inclusive of either spike Δ69-70 or ORF1a Δ3675-3677. To assess matrix effects on assay performance, both duplex assays were tested using samples of wild type or B.1.1.7 RNA diluted in RNA extract from wastewater previously tested to contain no SARS-CoV-2 RNA.

The performance of the duplex assays to estimate the ratio of VOCs in a mixed sample was tested in samples containing both wild type and B.1.1.7 RNA with the following proportions of variant: 0.01, 0.02, 0.10, 0.50, 0.90, 0.98, and 0.99, at a total target concentration of 100 gc/μl. Dilutions were performed in molecular grade water.

The limit of quantification (LOQ) was determined by measuring 10 replicates of 25, 30 and 40 gc/reaction using the wild type RNA. The LOQ was defined as the lowest value where the relative standard deviation is smaller than 25% (Zhu et al. 2020). For spike Δ69-70, the LOQ was at 40 gc/reaction, and for ORF1a Δ3675-3677 at 25 gc/reaction.

### Measuring VOCs in Wastewater

The drop-off RT-dPCR assays were applied to 32 raw influent (24-hour flow proportional composite) samples collected from the wastewater treatment plant Werdhölzli (serving approximately 450’000 people in Zurich, Switzerland) from 7 December 2020 to 26 March 2021. Samples were stored at 4°C for up to 8 days, shipped to Eawag (Duebendorf, Switzerland) for immediate concentration and RNA extraction, and stored at -80°C as RNA extracts for up to 4 months.

Sample concentration and RNA extraction followed the previously reported protocol (Huisman et al. 2021; Fernandez-Cassi et al., n.d.). In brief, raw influent (50 ml) was clarified by centrifugation or filtration (0.22 μM filter), then samples were concentrated using ultrafiltration (10 kDa Centricon Plus-70, Millipore, USA). RNA from viral concentrates was extracted using the QiaAmp Viral RNA MiniKit (Qiagen, USA), eluted in 80 μL, then, for samples after 14 January, further purified using OneStep PCR Inhibitor Removal Kits (Zymo Research) to reduce assay inhibition. RNA extracts were stored at -80°C for up to 4 months before quantification on dPCR. The dMIQE checklist is available in the Supplemental Material, Table S2 (Huggett et al. 2020).

### Data Analysis

The proportion of amplicons with deletions was calculated for each assay by accounting for the distribution of droplets with no, single, and double fluorescence (Supplemental Material, Text S1 and Figure S3). As previously described, we assume that the number *M*_*i*_ of virus RNA genome copies in a droplet *i* is Poisson distributed with mean *λ* = *Cv*, where *C* is the concentration of SARS-CoV-2 RNA in genome copies per volume and *v*is the droplet volume (Dube, Qin, and Ramakrishnan 2008; Basu 2017; Dorazio and Hunter 2015). In assays with multiple targets such as duplex assays, positive counts for each probe are typically assumed to be independent(Dube, Qin, and Ramakrishnan 2008; Whale A.S, Hugget J. F., Tzonev S. 2016; Milbury et al. 2014). In a drop-off assay, this assumption is flawed as the number of viral particles positive for the drop-off probe trapped in a partition cannot exceed the number of viral particles positive for the conserved probe. When the relative proportion of variant in the sample is very low or very high (which is a common use case), confidence intervals based on this assumption will be vastly flawed. Here, we further developed the statistical methodology for the duplex drop-off assay by dropping this problematic assumption. We assume that the number *N*_*i*_ of virus particles without the deletion (double positive droplets) in a droplet *i* is binomially distributed with sample size *M*_*i*_such that *r*_*wt*_ corresponds to the relative frequency of the wild type in the viral population.

Assuming independent and identically distributed fluorescence signals of droplets, the counts *X*_0_of double negative droplets (no virus RNA), *X*_1_of droplets positive for the universal probe but not for the deletion probe (variant RNA), and *x*_2_of double positive droplets (wild type RNA) follow a multinomial distribution with probabilities of success 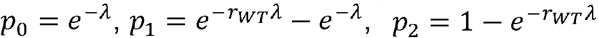 and sample size *n*_*tot*_ = *x*_0_ + *x*_1_ + *x*_3_corresponding to the total number of droplets (see Supplemental Material, Text S2 for details of derivation). From this likelihood, we find maximum likelihood estimators 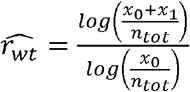 and 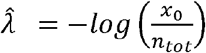. By invariance of the likelihood, the maximum likelihood estimator of the proportion of the mutant allele *r*_*mut*_ in the sample is 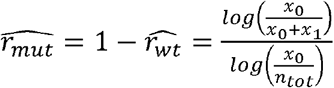.

The temporal increase in the proportion of amplicons with deletions in wastewater, indicative of VOCs, was compared to the temporal increase of the deletions and B.1.1.7 in clinical samples in Canton Zurich (region with 1.5 million inhabitants, Figure 2) and Switzerland (Supplemental Material, Figure S4). In analyzing clinical samples from Zurich and Switzerland, we assume that the proportion of clinical sample genomes that are B.1.1.7 or bear a signature mutation are binomially distributed, such that their empirical frequency is the maximum likelihood estimator for their prevalence in the population.

**Figure 2.**
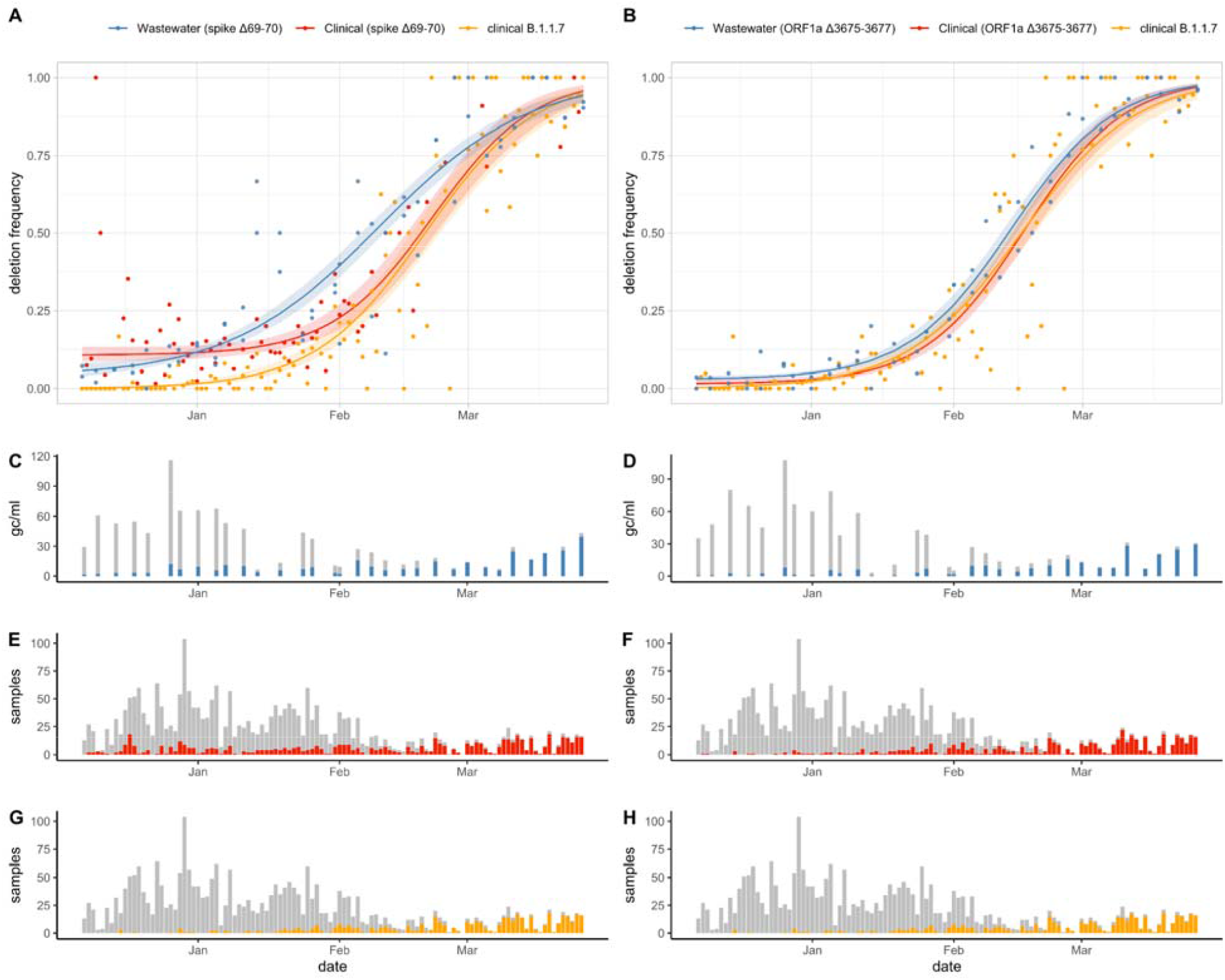
Proportion of (A) spike Δ69-70 and (B) ORF1a Δ3675-3677 in wastewater samples compared to Zurich clinical samples, along with the proportion of B.1.1.7 lineage in Zurich clinical samples. Fitted curves correspond to three-parametric logistic fits for the deletions in wastewater and clinical data and two-parametric logistic fit for the B.1.1.7 clinical data. Shaded areas correspond to 95% confidence bands. Concentration of SARS-CoV-2 RNA (grey) and concentration of deleted alleles (blue) for the (C) spike Δ69-70 and (D) ORF1a Δ3675-3677 in wastewater samples. Number of Zurich sequenced clinical samples (grey) and number of samples with the deleted allele (red) for the (E) spike Δ69-70 and (F) ORF1a Δ3675-3677. (G, H) Number of Zurich sequenced clinical samples (grey) and number of samples from the B.1.1.7 lineage (orange).

Following Chen et al. (Chen, Nadeau, Topolsky, Manceau, Huisman, Jablonski, et al. 2021), we assume that the relative frequency *r*(*t*) of a VOC follows a 2-parameter logistic (2PL) growth with rate *a* and inflection point *t*_0_, such that:

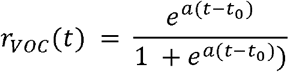

Notably, here and throughout, *t* is defined as days since 7 December 2020, the first day we included in analysis.

As some lineages with no reproductive fitness advantage already present in the region can share mutations with the VOC under investigation, we add a background constant prevalence parameter *c* when modelling the relative frequency *r*_*mut*_(*t*) of a mutation, such that it follows a 3-parametric logistic (3PL) curve (Baker and Kim 2004):

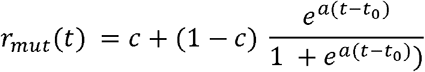

We assume each observation of the proportion of a mutation is an independent observation. Therefore, a best fit can be found by maximizing the joint (log-)likelihood of either the binomial counts in clinical data or the multinomial counts in wastewater data. The L-BFGS-B algorithm implemented in the R v4.0.5 package stats was used for the optimization and estimation of the Fisher information matrix. Wald confidence intervals for the parameter estimates were then computed, using the standard errors from quasi-binomial and quasi-multinomial distributions to account for over-/underdispersion (McCullagh and Nelder 2019; Held and Sabanés Bové 2014). The estimates of the growth rate parameter*a* were transformed along with their confidence intervals into transmission fitness advantage *f*_*d*_, assuming the discrete-time logistic growth model of Chen et al. and a generation time *g*= 4.8 days, such that *f*_*d*_ = *e*^*ag*^ − 1(Chen, Nadeau, Topolsky, Manceau, Huisman, Jablonski, et al. 2021). Wald confidence bands for the fitted values were calculated using the Delta method (Held, 2013) for the logit-transformed fitted values and then back transformed to the original scale, so as to ensure that they were restricted to the interval [0,1]. For each time series, both 3PL and 2PL were fit to the data and then compared by way of a likelihood ratio test based on quasi-likelihood (McCullagh and Nelder 2019), and the 3PL model was retained only if p < 0.05. The assumption of constant overdispersion of the quasi-likelihood models was checked graphically (Supplemental Material, Figure S5).

## RESULTS

### Benchmarking Drop-Off RT-dPCR Assay

Both drop-off assays targeting the spike Δ69-70 and the ORF1a Δ3675-3677 performed as expected in singleplex and duplex assays. Specifically, the universal probe was detected in singleplex and duplex assays when using RNA from both the B.1.1.7 VOC sample and the wild type strain. Similarly, the probe targeting only the wild type strain (deletion probe) was only detected in the wild type strain. Notably, when the assays were applied to the wild type strain RNA, the universal probe and the deletion probe reported nearly identical concentrations. In the spike Δ69-70 assay, the concentrations measured were 162.3 gc/μl for the universal probe and 162.8 gc/μl for the deletion probe. For the ORF1a Δ3675-3677 assay, the concentration measured was 155.2 gc/μl for both probes. When testing the ratio of the B.1.1.7 RNA to the wild type strain, both duplex assays performed as expected, with the exception of one replicate of the sample of the spike Δ69-70 assay with a variant to wild type RNA ratio of 1:50 (see Supplemental Material, Figure S6). In this sample, the variant RNA was not detected.

The assay performance was the same in wastewater matrix, with both the universal and deletion probes detectable in the wild type strain RNA and only the universal probe detectable in the VOC B.1.1.7 RNA. Specifically, concentrations were nearly identical for the wild type strain RNA for both the universal (43.3 gc/μl) and deletion (42.9 gc/μl) probes for the spike Δ69-70 assay, as well as for the universal (42.2 gc/μl) and deletion (42.2 gc/μl) probes for the ORF1a Δ3675-3677 assay.

### Measuring VOCs in Zurich Wastewater

Within the wastewater samples, concentrations reported by the universal probes (per ml wastewater) ranged from 2.0 gc/ml to 60.6 gc/ml for the spike Δ69-70 assay and from 0.50 gc/ml to 65.0 gc/ml for the ORF1a Δ3675-3677 assay (Figure 2). Concentrations of amplicons with deletions, estimated based on the number of single positive droplets, increased over time and ranged from 0.6 gc/ml to 20.4 gc/ml for spike Δ69-70 assay and from undetectable to 15.8 gc/ml for ORF1a Δ3675-3677 assay.

Wastewater detection of the deletions tracked the deletions in clinical samples from both Zurich and Switzerland (Figure 2 and Supplemental Material, Figure S4). Both deletions were detected in wastewater before the detection of B.1.1.7 in clinical samples (Figure 2), due to low frequencies of spike Δ69-70 and ORF1a Δ3675-3677 in non-B.1.1.7 clinical samples; spike Δ69-70 and ORF1a Δ3675-3677 were detectable in variants other than B.1.1.7, B.1.351, and P.1 at frequencies of 3.4% and 0.06% (out of 8’709 clinical samples) in Switzerland before December 24, 2020 (Jahn et al. 2021).

Accounting for these background prevalence rates in the logistic growth model, estimates for the growth rate, time to maximum growth, and transmission fitness advantage were generally consistent with estimates obtained from clinical samples across Switzerland (Table 2). Specifically, the growth rate estimate from wastewater was 0.06 [95% CI 0.06, 0.07] for spike Δ69-70 compared to an estimate of 0.09 [95% CI 0.07, 0.11] from Zurich clinical samples. Similarly, the growth rate estimate was 0.09 [95% CI 0.08, 0.09] for ORF1a Δ3675-3677 in wastewater compared to 0.09 [95% CI 0.08, 0.10] from Zurich clinical samples. Converting growth rates to transmission fitness advantage further highlighted the similarities, with wastewater estimates for spike Δ69-70 of 0.34 [95% CI 0.30, 0.39] and for ORF1a Δ3675-3677 of 0.53 [95% CI 0.39, 0.73], which are similar to estimates for both the deletions and for B.1.1.7 variant obtained from clinical samples, as demonstrated by overlapping confidence intervals (Table 2).

**Table 2.** Three parametric (3PL) logistic model parameter estimates for the prevalence of spike Δ69-70 and ORF1a Δ3675-3677 in wastewater data, Swiss clinical data and Zurich clinical data, as well as two-parametric (2PL) logistic model parameter estimates for the prevalence of B.1.1.7 variants in Swiss and Zurich clinical data. Values are maximum likelihood estimates and Wald 95% confidence intervals of the growth rate *a*, midpoint *t*_0_ (in days after December 07, 2020 and corresponding dates) and (in the case of 3PL) background prevalence *c*. Values for the rate parameter *a* are also shown transformed (along with their confidence intervals) into an estimate of the transmission fitness advantage *f*_*d*_ assuming the discrete-time growth model found in Chen et al. (Chen, Nadeau, Topolsky, Manceau, Huisman, Stadler, et al. 2021). 3PL models are shown only when the inclusion of a third parameter (Background Prevalence) is statistically significant (Supplemental Material, Table S2).

| Target | Sample Type | Growth Rate ($a$) | Time to Maximum Growth ($t_0$) | Background Prevalence ($c$) | Transmission Fitness Advantage ($f_d$) |
| --- | --- | --- | --- | --- | --- |
| B.1.1.7 | Switzerland (Clinical) | 0.07 [0.07, 0.08] | 66.1 [64.4, 67.8]<br>Feb 11 [9, 13] | 0 (fixed) | 0.42 [0.38, 0.46] |
|  | Zurich (Clinical) | 0.08 [0.07, 0.1] | 74.6 [70.0, 79.2]<br>Feb 20 [15, 24] | 0 (fixed) | 0.49 [0.38, 0.61] |
| spike $\Delta 69-70$ | Wastewater | 0.06 [0.06, 0.07] | 64.5 [62.0, 67.1]<br>Feb 10 [7, 12] | 0.04 [0.01, 0.06] | 0.34 [0.30, 0.39] |
|  | Switzerland (Clinical) | 0.07 [0.06, 0.08] | 65.5 [63.5, 67.6]<br>Feb 11 [8, 12] | 0.07 [0.04, 0.09] | 0.41 [0.36, 0.46] |
|  | Zurich (Clinical) | 0.09 [0.07, 0.11] | 76.3 [71.8, 80.9]<br>Feb 21 [17, 25] | 0.11 [0.07, 0.14] | 0.55 [0.39, 0.73] |
| ORF1a<br>$\Delta 3675-3677$ | Wastewater | 0.09 [0.08, 0.09] | 68.5 [67.4, 69.6]<br>Feb 14 [12, 14] | 0.03 [0.02, 0.04] | 0.53 [0.49, 0.57] |
|  | Switzerland (Clinical) | 0.09 [0.08, 0.11] | 68.7 [66.1, 71.2]<br>Feb 14 [11, 13] | 0.03 [0, 0.05] | 0.56 [0.45, 0.67] |
|  | Zurich (Clinical) | 0.09 [0.08, 0.1] | 71.3 [68.8, 73.7]<br>Feb 17 [15, 19] | 0.01 [0, 0.03] | 0.55 [0.46, 0.65] |

Time to maximum growth estimates were also generally similar when based on clinical or wastewater samples, falling within a range of less than two weeks in mid-February (10. - 21. February 2021). Here, estimates from Zurich clinical samples lagged behind both wastewater and Swiss clinical samples for both assays, with greater deviation for the spike Δ69-70 deletion (Table 2). Similarly, the estimated background prevalences *c* of the mutations were in general consistent between models based on the wastewater data and on the clinical data. The inclusion of a background prevalence parameter statistically significantly improved the fit in all models based on prevalence of deletions, but not in any model based on the prevalence of the B.1.1.7 in clinical data (Supplemental Material, Table S2).

## DISCUSSION

Drop-off RT-dPCR assays for targeted detection of signature mutations in wastewater show temporal trends consistent with sequencing data from clinical samples. By incorporating non-competitive probes on a single amplicon, proportions of the mutation in the population can be directly estimated within a single assay that is not biased by inter-assay variation in quantification. The analysis of wastewater in this study included 32 samples, which contrasts to 2’497 clinical samples collected in Canton Zurich and sequenced during the period of study (7 December 2020 to 26 March 2021). The demonstrated approach provides a method of tracking signature mutations within a community at lower cost and with faster turn around than clinical or wastewater sequencing, and can be adapted to screen for other signature mutations of other VOCs.

Temporal trends in signature mutations (here, ORF1a Δ3675-3677 and spike Δ69-70) of VOCs tracked in wastewater using dPCR have the potential to rapidly inform variant transmissibility. Growth rate and transmission fitness advantage estimates derived from wastewater were generally consistent to estimates derived from clinical samples, with overlapping 95% confidence intervals. Because dPCR sample analysis is faster than whole-genome sequencing, the demonstrated workflow could help to provide early insights into transmissibility of novel variants, once signature mutations are identified.

Notably, we found that the proportion of variants in wastewater increased earlier than in clinical samples in Zurich (Figure 2). The dates of the two types of samples represent distinct events, with the wastewater samples representing the 24-hour period within which the wastewater sample was collected, and the dates of the clinical samples representing the date a person visits a testing period. In wastewater samples, the maximum growth rate of ORF1a Δ3675-3677 occurred 2.8 days earlier than in clinical samples, and spike Δ69-70 occurred 11.8 days earlier than in clinical samples (Figure 2, Table 2). Previous work suggests Zurich wastewater signals for SARS-CoV-2 are at most 1 day earlier than clinical case data, based on the average delay between infection and date of testing in Zurich (8.1 days) compared to the estimated delay between infection and shedding of SARS-CoV-2 RNA in wastewater (7-11 days) (Huisman et al. 2021). One explanation for this discrepancy is that people infected with the B.1.1.7 variant shed substantially higher viral concentrations than people infected with non-B.1.1.7 variants (T. C. Jones et al. 2021). If more B.1.1.7 SARS-CoV-2 RNA is shed into wastewater per person infected as compared to infections with non-B.1.1.7 variants, then the proportion of variant amplicons in wastewater would be skewed towards B.1.1.7 and would not be an accurate estimate of the proportion of B.1.1.7 infections. Theoretically, wastewater-based estimates on frequency of VOCs in the population could be corrected to account for variant-specific differences in shedding, but this would require reliable estimates of shedding load profiles for each variant.

We applied the drop-off RT-dPCR assay here for signature mutations of B.1.1.7, P.1., and B.1.351, but the assay is readily adaptable to screen for other signature mutations beyond those presented here. Targeting signature deletions allows for the design of specific probes annealing to the deleted region to distinguish target variants from other lineages. However, probes providing specificity to other mutations, including single nucleotide polymorphisms, could be targeted by future drop-off assays. Examples include mutations that may influence not only transmissibility but also virulence and/or vaccine-induced antibody escape (Harvey et al. 2021).

Notably, wastewater-based estimation of the proportion of signature mutations in communities requires sufficient concentrations of SARS-CoV-2 RNA such that relative proportions of target mutations can be quantified. Despite wastewater concentrations of the deletion amplicons being often below the limit of quantification (defined as the lowest concentration with a coefficient of variation of less than or equal to 25%), transmission fitness estimates remain aligned with estimates from clinical samples. This finding suggests low SARS-CoV-2 RNA concentrations in wastewater may nevertheless be sufficient to infer epidemiological data when analyzed as a time series.

Drop-off RT-dPCR assays provide an opportunity for rapid detection and quantitative estimates of VOCs within communities, offering epidemiological insights into their introduction and transmission. Such assays offer complementary tools to sequencing analyses of VOCs which may be useful for large-scale programs in wastewater-based epidemiology of SARS-CoV-2 such as the U.S. National Sewer Surveillance System and the EU Sewer Sentinel Surveillance Network.

## Data Availability

The data obtained from wastewater samples are available by contacting the authors, and will be archived in a public database upon article publication following peer review. The data on clinical samples were accessed by the publicly available covSPECTRUM database (https://cov-spectrum.ethz.ch).

## AUTHOR CONTRIBUTIONS

LC, TK, CO, and TRJ, conceived the study; DD and NB developed and applied statistical and analytical methods; LC, TS, AJD, PG, AK, ES, XFC, TK, CO, and TRJ developed experimental protocols and performed wastewater sampling; XFC, NB, TK, CO, and TRJ supervised the study and secured funding; LC, DD, and TRJ wrote the original draft; all authors reviewed and approved the final manuscript.

## ACKNOWLEDGEMENTS

We thank the operators of the Zurich WWTPs for providing samples. Funding provided through the Swiss National Science foundation (Special Call on Coronaviruses (31CA30_196538), Federal Office of Public Health, Eawag and EPFL discretionary funding. XFC was a fellow of the European Union’s Horizon 2020 research and innovation programme under the Marie Skłodowska–Curie Grant Agreement No. 754462.

