## Supplemental Material for "Inferring Transmission Fitness Advantage of SARS-CoV-2 Variants of Concern in Wastewater Using Digital PCR"

#### Supplemental Material Contains:

**Table S1.** Composition of mastermix for both assays

**Figure S1.** Impact of wastewater dilutions on concentrations of SARS-CoV-2 RNA per reaction volume

**Table S2.** dMIQE guidelines

**Figure S2.** Scatterplot of positive and negative control for both assays

**Text S1.** Analysis of droplets in 2D scatter plot

**Figure S3.** Analysis of droplets in 2D scatter plot

**Text S2.** Supplementary Statistical Analysis

**Figure S4.** Proportion of spike Δ69-70 and ORF1a Δ3675-3677 in wastewater samples compared to clinical samples from throughout Switzerland

**Figure S5.** Overdispersion vs. Sample Size for the 2PL and 3PL models of the logistic growth of the spike Δ69-70 and ORF1a Δ3675-3677 deletions in wastewater and B.1.1.7 in clinical samples

**Figure S6.** Proportion of amplicons containing either the spike Δ69-70 or ORF1a Δ3675-3677 deletions relative to expectation

**Table S2.** Three parametric (3PL) and two-parametric (2PL) logistic model parameter estimates for the prevalence of spike Δ69-70 and ORF1a Δ3675-3677 in wastewater data, Swiss clinical data and Zurich clinical data

**Table S1.** Composition of mastermix for both assays, spike Δ69-70 and ORF1a Δ3675-3677.

|  | **spike Δ69-70** | |  | **ORF1a Δ3675-3677** | |
| --- | --- | --- | --- | --- | --- |
|  | Final concentration | Volume 1 sample (µL) |  | Final concentration | Volume 1 sample (µL) |
| RNAse Free Water | - | 3.915 |  | - | 3.78 |
| qScript XLT One-Step RT-qPCR ToughMix 2x | 1x | 13.5 |  | 1x | 13.5 |
| Fluorescein 1 µM | 100 nM | 2.7 |  | 100 nM | 2.7 |
| Forward Primer (20 µM) | 0.4 µM | 0.54 |  | 0.4 µM | 0.54 |
| Reverse Primer (20 µM) | 0.4 µM | 0.54 |  | 0.4 µM | 0.54 |
| Deletion Probe (20 µM) | 0.1 µM | 0.135 |  | 0.2 µM | 0.27 |
| Universal Probe (20 µM) | 0.2 µM | 0.27 |  | 0.2 µM | 0.27 |
| Total |  | 21.6 |  |  | 21.6 |

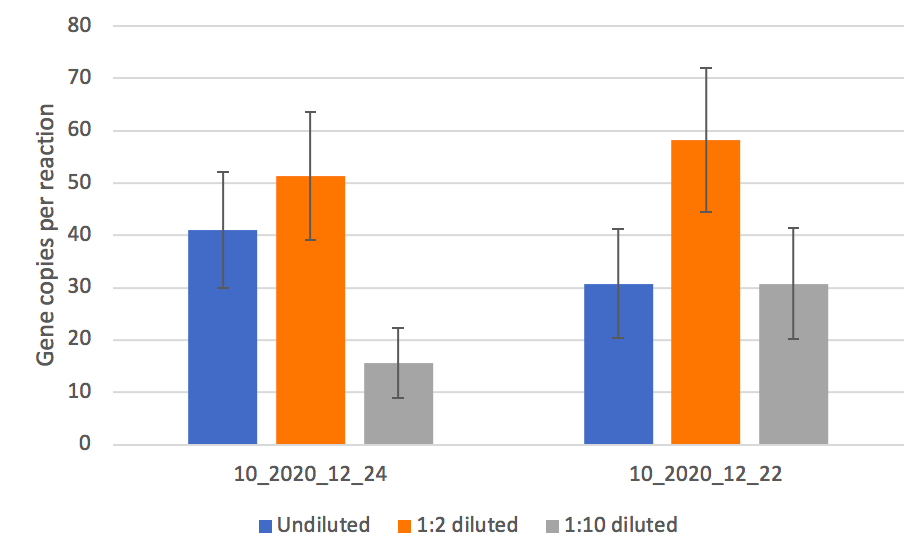

**Figure S1.** Two wastewater samples (10_2020_12_24 and 10_2020_12_22, referring to samples from December 22 and 24 in 2020 from wastewater treatment plant Werdhölzli) were measured using the spike Δ69-70 assay in three dilutions: undiluted (5 µl RNA), 1:2 diluted (2.5 µl RNA) and 1:10 diluted (0.5 µl RNA). Each dilution was measured in duplicate and the average was plotted. Error bars indicate the 95% confidence interval. The ordinate shows gene copies per 25 µl reaction, indicating that a 1:2 dilution results in the highest number of detectable genes per reaction.

**Table S2.** dMIQE guidelines

| **ITEM TO CHECK** | **PROVIDED** | **COMMENT** |
| --- | --- | --- |
| **Column1** | **Y/N** | **Column2** |
| **1. SPECIMEN** |  |  |
| Detailed description of specimen type and numbers | **Y** | 32 RNA extractions from waste water treatment plant Werdhölzli in Switzerland after first treatment step (rake) - 24-hour flow proportional composite samples. Samples range from December 7th to March 26th. Calibration of ddPCR assay was performed using RNA from the Wuhan-Hu-1 SARS-CoV-2 strain (wild type RNA) and RNA from the B.1.1.7 SARS-CoV-2 strain (variant RNA) - both provided by Tobias Schindler, TPH. |
| Sampling procedure (including time to storage) | **Y** | 24-hour flow proportional composite samples |
| Sample aliquotation, storage conditions and duration | **Y** | Stored at 4°C for up to 8 days, shipped to Eawag for concentration and RNA extraction. |
| **2. NUCLEIC ACID EXTRACTION** |  |  |
| Description of extraction method including amount of sample processed | **Y** | Raw influent (50 ml) was clarified by centrifugation or filtration (0.22 μM filter), then samples were concentrated using ultrafiltration (10 kDa Centricon Plus-70, Millipore, USA). RNA from viral concentrates was extracted using the QiaAmp Viral RNA MiniKit (Qiagen, USA), eluted in 80 μL, then, for samples after 14 January, further purified using OneStep PCR Inhibitor Removal Kits (Zymo Research, CND 6030) to reduce assay inhibition. |
| Volume of solvent used to elute/resuspend extract | **Y** | 80 μL eluent volume |
| Number of extraction replicates | **Y** | Each sample was extracted once (no replicates). |
| Extraction blanks included? | **Y** | Extraction negative controls were included for every 10-14 samples. |
| **3. NUCLEIC ACID ASSESSMENT AND STORAGE** |  |  |
| Method to evaluate quality of nucleic acids | **N** | Not done |
| Method to evaluate quantity of nucleic acids (including molecular weight and calculations when using mass) | **N** | Not done |
| Storage conditions: temperature, concentration, duration, buffer, aliquots | **Y** | RNA extracts were stored at -80°C for up to 4 months before quantification on dPCR. |
| Clear description of dilution steps used to prepare working DNA solution | **Y** | RNA extracts from wastewater were diluted 1:2 (1 part RNA extract with 1 part RNAse free water) to maximize the number of gene copies per reaction, which is influenced by sample inhibition. |
| **4. NUCLEIC ACID MODIFICATION** |  |  |
| Template modification (digestion, sonication, pre-amplification, bisulphite etc.) | **N** | Not applicable |
| Details of repurification following modification if performed | **N** | Not applicable |
| **5. REVERSE TRANSCRIPTION** |  |  |
| cDNA priming method and concentration | **N** | Not applicable |
| One or two step protocol (include reaction details for two step) | **Y** | One-step protocol |
| Amount of RNA added per reaction | **Y** | 5 μl RNA extract 1:2 diluted per 25 μl PCR assay |
| Detailed reaction components and conditions | **Y** | See below (7. dPCR PROTOCOL) |
| Estimated copies measured with and without addition of RT* | **N** | Not done |
| Manufacturer of reagents used with catalogue and lot numbers | **Y** | See below (7. dPCR PROTOCOL) |
| Storage of cDNA: temperature, concentration, duration, buffer and aliquots | **N** | Not applicable: cDNA was used directly for PCR step |
| **6. dPCR OLIGONUCLEOTIDES DESIGN AND TARGET INFORMATION** |  |  |
| Sequence accession number or official gene symbol | **Y** | spike Δ69-70 and ORF1a Δ3675-3677 |
| Method (software) used for design and *in silico* verification | **Y** | IDT oligo analyzer |
| Location of amplicon | **Y** | spike Δ69-70: 21'710-21'817 bp, ORF1a Δ3675-3677: 11'229-11'356 bp (based on NC_045512.2:21563-25384 Severe acute respiratory syndrome coronavirus 2 isolate Wuhan-Hu-1, complete genome) |
| Amplicon length | **Y** | spike Δ69-70: 108 bp, ORF1a Δ3675-3677: 128 bp |
| Primer and probe sequences (or amplicon context sequence)** | **Y** | spike Δ69-70: Forward primer 5'-TCAACTCAGGACTTGTTCTTACCT-3’, Reverse primer 5'-TGGTAGGACAGGGTTATCAAAC-3', Deletion probe 5'-Cy5-TTCCATGCTATACATGTCTCTGGGA-BHQ2-3',  Universal probe 5'-HEX-CCAATGGTACTAAGAG-MGBQ530-3'  ORF1a Δ3675-3677: Forward primer 5'-TGCCTGCTAGTTGGGTGATG-3', Reverse primer 5'-TGCTGTCATAAGGATTAGTAACACT-3', Deletion probe  5'-HEX-GTTTGTCTGGTTTTAAGCTAAAAGACTGTG-BHQ1-3',  Universal probe 5'-Cy5-CGTATTATGACATGGTTGGATATGGTTGAT-BHQ2-3' |
| Location and identity of any modifications | **Y** | Reporter and Quencher of probes see above |
| Manufacturer of oligonucleotides | **Y** | Microsynth AG in Balgach Switzerland |
| **7. dPCR PROTOCOL** |  |  |
| Manufacturer of dPCR instrument and instrument model | **Y** | Stilla Technologies, Naica System |
| Buffer/kit manufacturer with catalogue and lot number | **Y** | qScript XLT 1-Step RT-PCR Kit (QuantaBio 95132-500, Lot number 66171338 and 66172618 and QuantaBio 95132-02K, Lot number 661726619) |
| Primer and probe concentration | **Y** | Final concentrations:  spike Δ69-70: Primers 0.4 μM, Deletion probe 0.1 μM, Universal probe 0.2 μM ORF1a Δ3675-3677: Primers 0.4 μM, Probes 0.2 μM |
| Pre-reaction volume and composition (incl. amount of template and if restriction enzyme added) | **Y** | Total volume of 27 μl: 13.5 μl qScript, 2.7 μl Fluorescein (final concentration 100 nM), 0.54 μl forward primer, 0.54 μl reverse primer, 0.27 μl deletion probe (0.135 μl deletion probe for spike Δ69-70 assay), 0.27 μl universal probe, water up to a final volume of 21.6 μl, 5.4 μl RNA template. 25 μl were loaded on the chip. |
| Template treatment (initial heating or chemical denaturation) | **Y** | RNA extracts from wastewater were diluted 1:2 to maximize the number of gene copies per reaction, which is influenced by sample inhibition (2.7 μl RNA extract and 2.7 μl water) |
| Polymerase identity and concentration, Mg++ and dNTP concentrations*** | **Y** | Based on manufacturer's preparation (not known): QuantaBio 95132-500 and QuantaBio 95132-02K |
| Complete thermocycling parameters | **Y** | Partitioning 12 min/40°C //  Reverse transcription 30 min/55°C // Enzyme activation 1 min/95°C //  40 cycles 10 sec/95°C and 30 sec/55°C |
| **8. ASSAY VALIDATION** |  |  |
| Details of optimisation performed | **Y** | Procedure for both deletions (spike Δ69-70 and ORF1a Δ3675-3677):  1. Singleplex against wild type and variant RNA individually  2. Duplex against wild type and variant RNA individually  3. Different ratios of wild type and variant RNA in duplex assays  4. LOQ measurements using 25, 30 and 40 copies/reaction |
| Analytical specificity (vs. related sequences) and limit of blank (LOB) | **N** | not done |
| Analytical sensitivity/LoD and how this was evaluated | **Y** | Negative control has less than 3 positive droplets, otherwise the experiment has to be repeated. A sample is positive with at least 3 positive droplets. |
| Testing for inhibitors (from biological matrix/extraction) | **Y** | 2 samples (10_2020_12_22 and 10_2020_12_24) were measured using the spike Δ69-70 assay in different dilutions: undiluted, 1:2 diluted and 1:10 diluted. 1:2 dilution gave the highest gene copy numbers per reaction. For details see Supplemental Material Figure S1. |
| **9. DATA ANALYSIS** |  |  |
| Description of dPCR experimental design | **Y** | Measuring wastewater samples over time targeting two different SARS-CoV-2 deletions: spike Δ69-70 and ORF1a Δ3675-3677 |
| Comprehensive details negative and positive of controls (whether applied for QC or for estimation of error) | **Y** | In each run, a negative sample (water as RNA template) and a positive sample (wild type RNA in a known concentration) were included. |
| Partition classification method (thresholding) | **Y** | In each run, polygons were manually drawn according to the description in Supplemental Material Text S1. Basically 3 polygons were drawn: a negative, a single positive and a double positive polygon according to the Technical Note from Stilla. |
| Examples of positive and negative experimental results (including fluorescence plots in supplemental material) | **Y** | See Supplemental Material, Figure S2. |
| Description of technical replication | **Y** | Each sample was measured in duplicates. |
| Repeatability (intra-experiment variation) | **Y** | 10 replicates of a known concentration (25 copies/reaction) were run on both assays. The average (and relative standard deviation) for each assay are given as: 22.65 copies/reaction (27.61%) for spike Δ69-70 and 27.38 copies/reaction (22.92%) for ORF1a Δ3675-3677 |
| Reproducibility (inter-experiment/user/lab etc. variation ) | **Y** | All experiments were performed in the same lab, on the same instruments by the same person, but on different days. Performance of positive control at 1'000 copies/reaction (n=6 for each assay) for spike Δ69-70 showed a relative standard deviation of 4.16% and for ORF1a Δ3675-3677 a relative standard deviation of 4.43%. |
| Number of partitions measured (average and standard deviation ) | **Y** | spike Δ69-70: 25’205 droplets with a standard deviation of 2’289 droplets (n=162)  ORF1a Δ3675-3677: 25’716 droplets with a standard deviation of 3’103 droplets (n=143) |
| Partition volume | **Y** | Diameter of droplets when using qScript on Stilla is 101.53 μm. This equals a spheric volume of 548001.542 μm^3 or 0.000548 μl. |
| Copies per partition (λ or equivalent ) (average and standard deviation) | **Y** | spike Δ69-70: λ equals 0.01177 with a relative uncertainty of 11.37%  ORF1a Δ3675-3677: λ equals 0.01389 with a relative uncertainty of 10.38% |
| dPCR analysis program (source, version) | **Y** | CrystalReader and CrystalMiner 2.4.0.3 |
| Description of normalisation method | **N** | not applicable |
| Statistical methods used for analysis | **Y** | see main text |
| Data transparency | raw data uploaded to online repository with ID: | DOI available on acceptance |

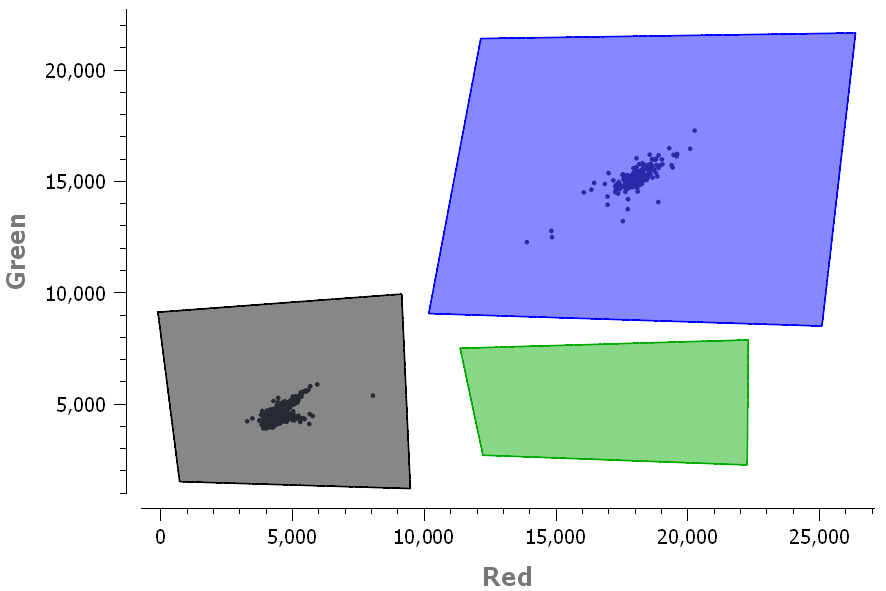

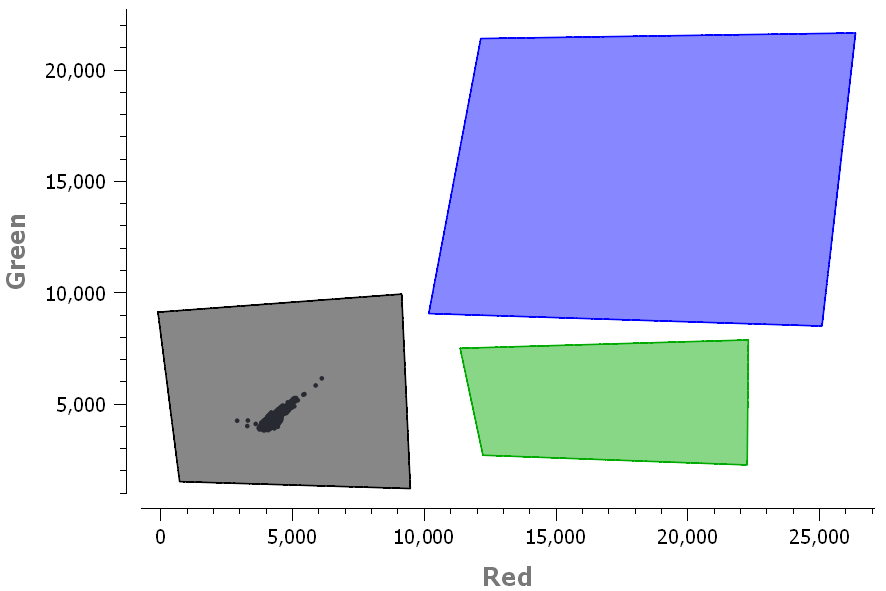

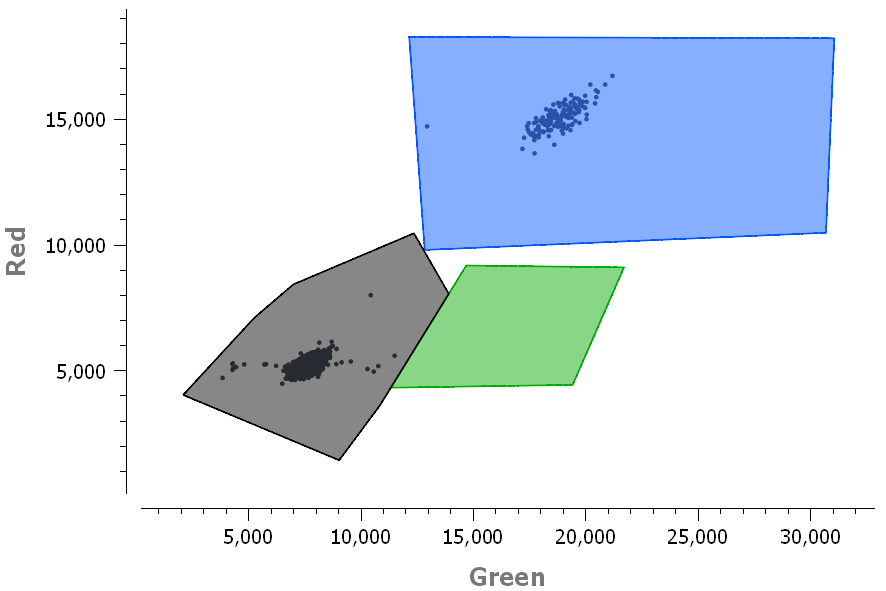

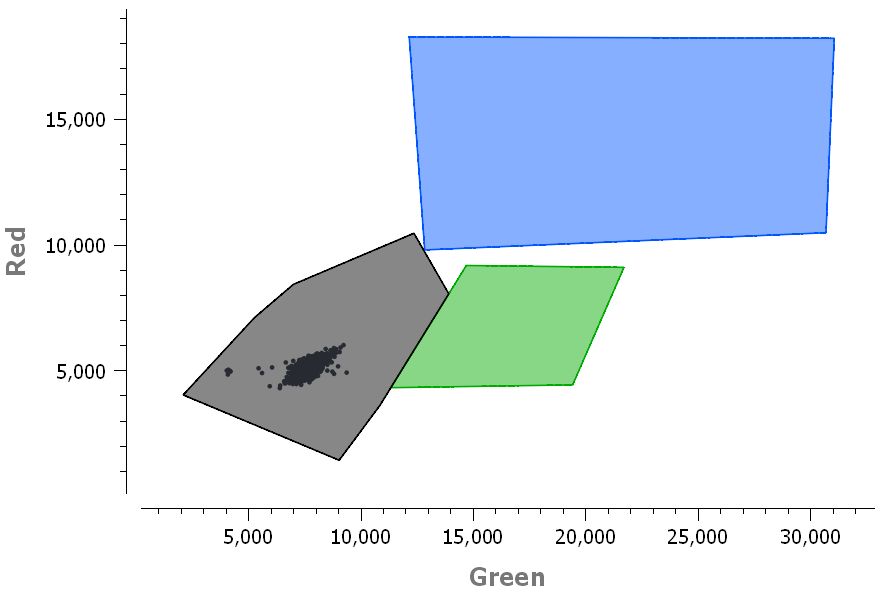

**Figure S2.** 2D scatter plot of digital PCR droplet fluorescence of the drop-off dPCR assays for positive (top left, bottom left) and negative (top right, bottom right) controls for both the spike Δ69-70 (top left, top right) and ORF1a Δ3675-3677 (bottom left, bottom right) assays. Positive controls included SARS-CoV-2 wild type RNA at a target concentration of 100 gc/reaction as the template. Negative controls included molecular grade water as the template. Polygons in each image represent non-fluorescing negative droplets (black), single positive droplets (green) and double positive droplets (blue). Because SARS-CoV-2 wild type RNA does not contain the target deletions, the sample is double fluorescing (blue polygon).

**Text S1.** Analysis of droplets in 2D scatter plot

Droplets were analyzed using the 2D scatter plot. Thresholds were set by drawing polygons according to the technical note of Stilla. In short, three polygons are required: one for the negative droplets, one for the single positive droplets (variants) and one for the double positive droplets (wild type). Using the Population Editor of Stilla, the negative droplets were then associated with the single positive droplets, following manufacturer recommendations (<https://www.stillatechnologies.com/how-to-quantify-drop-off-digital-pcr-assays-with-crystal-miner/>, accessed 24.07.2021).

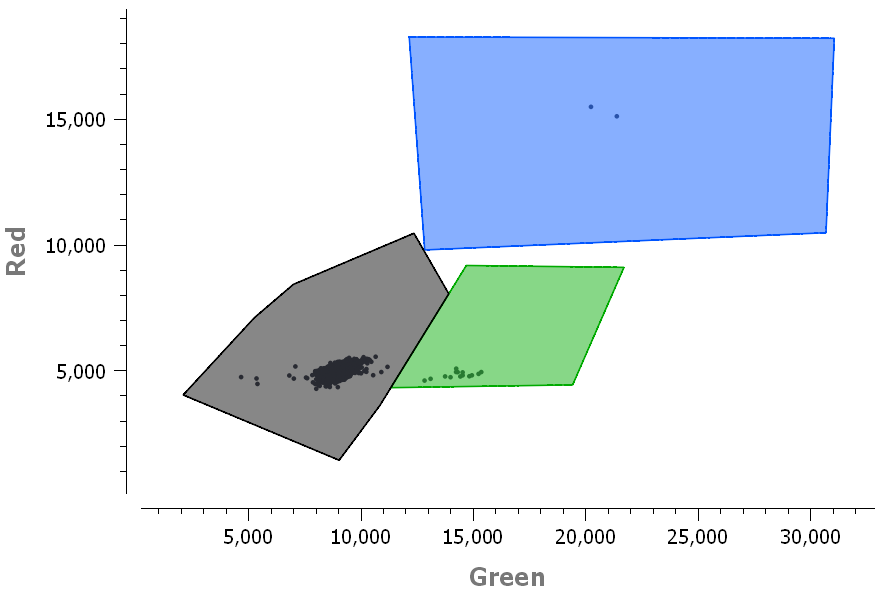

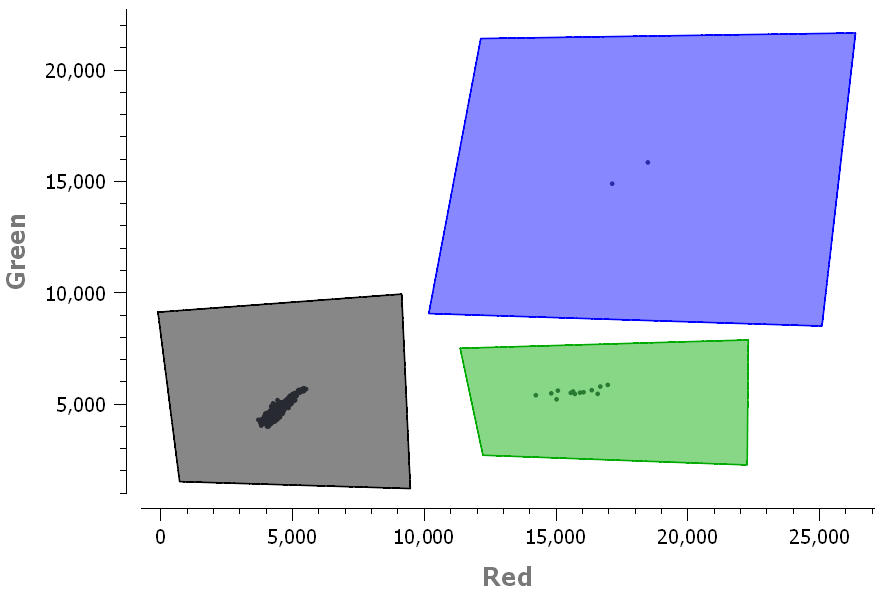

**Figure S3.** 2D scatter plot of digital PCR droplet fluorescence. Polygons were drawn around single positive (green polygon), double positive (blue polygon) and negative droplets (black polygon). Images represent sample 10_2021_03_01, which is the sample collected from Zurich wastewater treatment plant on March 03, 2021 at 1:2 dilution (replicate b) for the spike Δ69-70 (top) and ORF1a Δ3675-3677 (bottom) assay.

**Text S2.** Supplementary Statistical Analysis

##### Likelihood for dPCR data based on independent Binomial Poisson statistics

Let $X_{0}, X_{1}, X_{2}$ be the counts of partitions that are negative, positive for one probe, and positive for both probes, respectively. Let us denote by $\lambda$ the normalized viral RNA probe (universal probe) concentration in solution and by $r$ the proportion of the dropout probe (deletion probe). If we assume independent counts, where positivity of the droplets is as usual the outcome of Poisson statistics,

$$X_{2} \sim Binomial(n=x_{0}+x_{1}+x_{2}, p=1-e^{-r\lambda}) , X_{1}+X_{2} \sim Binomial(n=x_{0}+x_{1}+x_{2}, p=1-e^{-\lambda})$$

then we can then form the likelihood

$$L(r,\lambda)\propto(e^{-\lambda})^{x_{0}}(1-e^{-r\lambda})^{x_{2}}(e^{-r\lambda})^{x_{0}+x_{1}}(1-e^{-\lambda})^{x_{1}+x_{2}}$$

and the log-likelihood kernel

$$l(r,\lambda)=-x_{0}\lambda-(x_{0}+x_{1})r\lambda+x_{2}log(1-e^{-r\lambda})+(x_{1}+x_{2})log(1-e^{-\lambda})$$

We can find MLEs for both parameters:

$$\hat{\lambda} =-log\left( \frac{x_{0}}{x_{0}+x_{1}+x_{2}} \right)=-log\left( 1-\frac{x_{1}+x_{2}}{x_{0}+x_{1}+x_{2}} \right)$$

$$\hat{r}=\frac{log\left( \frac{x_{0}+x_{1}}{n} \right)}{log\left( \frac{x_{0}}{n} \right)}$$

##### Likelihood for dPCR data based on dependent counts

We can drop the assumption of droplet counts generated by independent Binomial Poisson statistics. We consider that droplets have a zero probability of being positive for the dropout probe and negative for the generic probe simultaneously. The counts $X_{0}, X_{1}, X_{2}$respectively of double negative partition, positive for the dropout probe, positive for generic probes are then assumed to follow a Multinomial distribution. Let us again denote by $\lambda$ the normalized viral RNA probe concentration in solution and by $r$the proportion of the dropout probe. We will first derive the probability parameters of the multinomial distribution.

Let us denote by $M_{i}$ the number of target particles in cell $i$ and assume that it is Poisson distributed,

$$M_{i}\sim Poisson(\lambda)$$

Each of these particles can be positive or negative for the dropout probe, such that the count $N_{i}$ of positive dropout probes in the cell $i$ is binomially distributed knowing the counts $M_{i}$,

$$N_{i} \mid M_{i}\sim Binomial(r, m_{i})$$

It can be show that the marginal distribution of $N_{i}$is also Poisson,

$$N_{i}\sim Poisson(r\lambda)$$

The probability $p_{0}$of double negative droplets is therefore

$p_{0}=Pr(N_{i}=0,M_{i}=0)=Pr(N_{i}=0 \mid M_{i}=0)Pr(M_{i}=0)=1\cdot e^{-\lambda}$

The probability $p_{1}$of single positive droplets can be found as

$$p_{1}=Pr(N_{i}=0,M_{i}\geq1) =Pr(M_{i}\geq1 \mid N_{i}=0)Pr(N_{i}=0) =\left( 1 - Pr(M_{i}=0 \mid N_{i}=0) \right)Pr(N_{i}=0)$$

$$=\left( 1-\frac{Pr(N_{i}=0,M_{i}=0)}{Pr(N_{i}=0)} \right)Pr(N_{i}=0) =Pr(N_{i}=0) - Pr(N_{i}=0, M_{i}=0)=e^{-r\lambda} - e^{-\lambda}$$

And we can find the probability of the last partition by subtracting the probabilities of the other partitions from unity:

$$p_{2}=Pr(N_{i}\geq1, M_{i}\geq1)=1 - \left( e^{-\lambda}+e^{-r\lambda} - e^{-\lambda} \right)=1 - e^{-r\lambda}$$

Assuming that all cells are independent and identically distributed, we have, for the total counts of droplets

$${(X}_{0}, X_{1}, X_{2}) \sim Multinomial(p_{0}=e^{-\lambda}, p_{1}=e^{-r\lambda} - e^{-\lambda}, p_{2}=1 - e^{-r\lambda}, n_{tot})$$

We can then form the likelihood for the parameters $\lambda,r$ as

$$L(\lambda,r)=\frac{n_{tot}!}{x_{0}!x_{1}!x_{2}!}\left( e^{-\lambda} \right)^{x_{0}}\left( e^{-r\lambda}-e^{-\lambda} \right)^{x_{1}}\left( 1-e^{-r\lambda} \right)^{x_{2}}$$

such that we have the log-likelihood kernel

$$\mathcal{l(}\lambda,r)=-x_{0}\lambda+x_{1}log\left( e^{-r\lambda}-e^{-\lambda} \right)+x_{2}log\left( 1-e^{-r\lambda} \right)$$

The MLEs are

$$\hat{r}={argzero}_{r}\frac{\partial\mathcal{l(}\lambda,r)}{\partial r}={argzero}_{r}\left\{ -\frac{\lambda x_{1}e^{-\lambda r}}{e^{-\lambda r}-e^{-\lambda}}+\frac{\lambda x_{2}e^{-\lambda r}}{1-e^{-\lambda r}} \right\}=\frac{\lambda+log\left( x_{1}+x_{2} \right)-log\left( x_{1}e^{\lambda}+x_{2} \right)}{\lambda}$$

and

$$\hat{\lambda}={argzero}_{\lambda}\frac{\partial\mathcal{l(}\lambda,r)}{\partial\lambda}={argzero}_{\lambda}\left\{ \frac{rx_{2}e^{-\lambda r}}{1-e^{-\lambda r}}-x_{0}+\frac{x_{1}\left( -re^{-\lambda r}+e^{-\lambda} \right)}{e^{-\lambda r}-e^{-\lambda}} \right\}$$

Substituting $\hat{r}$ in the expression for $\hat{\lambda}$ and then vice versa yields the same MLEs as when assuming independent Binomials:

$$\hat{\lambda}=-log\left( \frac{x_{0}}{x_{0}+x_{1}+x_{2}} \right)=-log\left( 1-\frac{x_{1}+x_{2}}{x_{0}+x_{1}+x_{2}} \right)$$

$$\hat{r}=\frac{log\left( \frac{x_{0}+x_{1}}{n} \right)}{log\left( \frac{x_{0}}{n} \right)}$$

Although the MLEs are the same as in the independent Binomials case, the likelihoods have much different forms (and different supports). To do simultaneous inference on $\lambda$ and $r$, we can build $1-\alpha$ confidence regions for $(r^,\lambda^)$ by inverting a likelihood ratio test and finding the set of points

$$\left\{ (r,\lambda\mathcal{): l(}r,\lambda\mathcal{)=l(}\hat{r},\hat{\lambda})-\frac{1}{2}\chi_{1,1-\alpha}^{2} \right\}$$

Where $\chi_{1,1-\alpha}^{2}$ is the $1-\alpha$ quantile of the chi-squared distribution with 1 degree of freedom. The above expression cannot in general be solved exactly, but it can be approximated arbitrarily well.

Since we are mainly interested in making inference for $r$, we can treat $\lambda$as a nuisance parameter and we can define the profile log-likelihood for $r$ as

$$\mathcal{l}_{p}(r) =\max_{\lambda}\mathcal{l(}r,\lambda\mathcal{)=l(}r,\hat{\lambda}(r)) =-\hat{\lambda}(r)x_{0}+x_{1}log\left( e^{-\hat{\lambda}(r)r}-e^{-\hat{\lambda}(r)} \right)+x_{2}log\left( 1-e^{-\hat{\lambda}(r)r} \right)$$

and the estimated profile log-likelihood for $r$ as

$$\mathcal{l}_{e}(r\mathcal{) =l(}r,\hat{\lambda}) =x_{0}log\left( \frac{x_{0}}{x_{0}+x_{1}+x_{2}} \right)+x_{1}log\left( -\frac{x_{0}}{x_{0}+x_{1}+x_{2}}+e^{rlog\left( \frac{x_{0}}{x_{0}+x_{1}+x_{2}} \right)} \right)+x_{2}log\left( 1-e^{rlog\left( \frac{x_{0}}{x_{0}+x_{1}+x_{2}} \right)} \right)$$

From these expressions, we can find asymptotic $(1-\alpha)$ confidence intervals for $r^$ based on the (estimated) profile likelihood ratio, by finding the strictly positive values of $r$ that solve:

$$2\left( \mathcal{l}_{p}(r)-\mathcal{l}_{p}(\hat{r}) \right)+\chi_{1,1-\alpha}^{2}=0$$

Where $\chi_{1,1-\alpha}^{2}$ is the $1-\alpha$ quantile of a chi-square distribution with 1 degree of freedom. These intervals will be nicely behaved (for example, they do not include negative values), but they require numerical root-finding of the expression above. The interval based on the profile likelihood better takes into account uncertainty in $\hat{\lambda}$, but is more difficult to compute as it requires optimization in terms of $\lambda$in each iteration of the root-finding algorithm.

##### Fitting a logistic curve to the data

We consider measures in a time series independent and distributed according to the same family: (quasi)binomial for clinical data and (quasi)multinomial for dPCR data. For the (quasi)binomial data, the success probability parameter is taken as time varying $r(t;a,b,c)$ taking the form of a 2PL or 3PL. For the (quasi)multinomial data, ${\lambda(t)}$is considered as a nuisance parameter, and we optimize the estimated profile log likelihood:

$$\mathcal{l}_{e}(a,b,c;t_{1:T})=\mathcal{l}(a,b,c,\hat{\lambda}_{1:T};t_{1:T}) =\sum_{i=1}^{T} \mathcal{l(}r(t_{i};a,b,c),\hat{\lambda}_{i})$$

Optimization and estimation of the Fisher information matrix is carried out using the L-BFGS-B algorithm implemented in the R v4.0.5 package stats. For the 3PL, the process is very sensitive to starting values (Baker and Kim 2004). Wald confidence intervals for parameters $a, b, c$ can be found using the estimate of the observed Fisher information matrix:

$${se}^{2}\left( [\hat{a},\hat{b},\hat{c}]_{i} \right)=\sqrt{[I(\hat{a},\hat{b},\hat{c})^{-1}]_{ii}}$$

Here, we use an inflated version of the inverse Fisher information matrix according to the moment estimated over-/underdispersion factor from a quasibinomial or quasimultinomial quasilikelihood (McCullagh and Nelder 2019), which amount to the average ratios of square deviations from the expected counts scaled by the variances from binomial or multinomial models. Standard errors for parameters $a, b, c$ can then be translated into standard errors for the function $r(t)$ by the delta method:

$${se}^{2}\left( r(t;a,b,c)) \right)=J_{r}(\hat{a},\hat{b},\hat{c})I(\hat{a},\hat{b},\hat{c})^{-1}J_{r}(\hat{a},\hat{b},\hat{c})^{T}$$

where $J_{r}(a,b,c)$ is the Jacobian

$$J_{r}(a,b,c) =\left[ \frac{\partial r(t;a,b,c)}{\partial a},\frac{\partial r(t;a,b,c)}{\partial b},\frac{\partial r(t;a,b,c)}{\partial c} \right]$$

But we here instead choose to make confidence bands for the logit transform of $r(t)$,

$$\psi(r)=log\frac{r(t)}{1-r(t)}$$

and we then back-transform them to the original scale with

$$r(\psi)=\frac{1}{e^{-\psi}+1}$$

This process will ensure that values are confined to the interval [0,1]. We construct the confidence bands for $\psi$ using the delta method,

$${se}^{2}\left( \psi(t;a,b,c)) \right)=J_{\psi}(\hat{a},\hat{b},\hat{c})I(\hat{a},\hat{b},\hat{c})^{-1}J_{\psi}(\hat{a},\hat{b},\hat{c})^{T}$$

where $J_{\psi}(a,b,c)$ is the Jacobian

$$J_{\psi}(a,b,c) =\left[ \frac{\partial\psi(t;a,b,c)}{\partial a},\frac{\partial\psi(t;a,b,c)}{\partial b},\frac{\partial\psi(t;a,b,c)}{\partial c} \right]$$

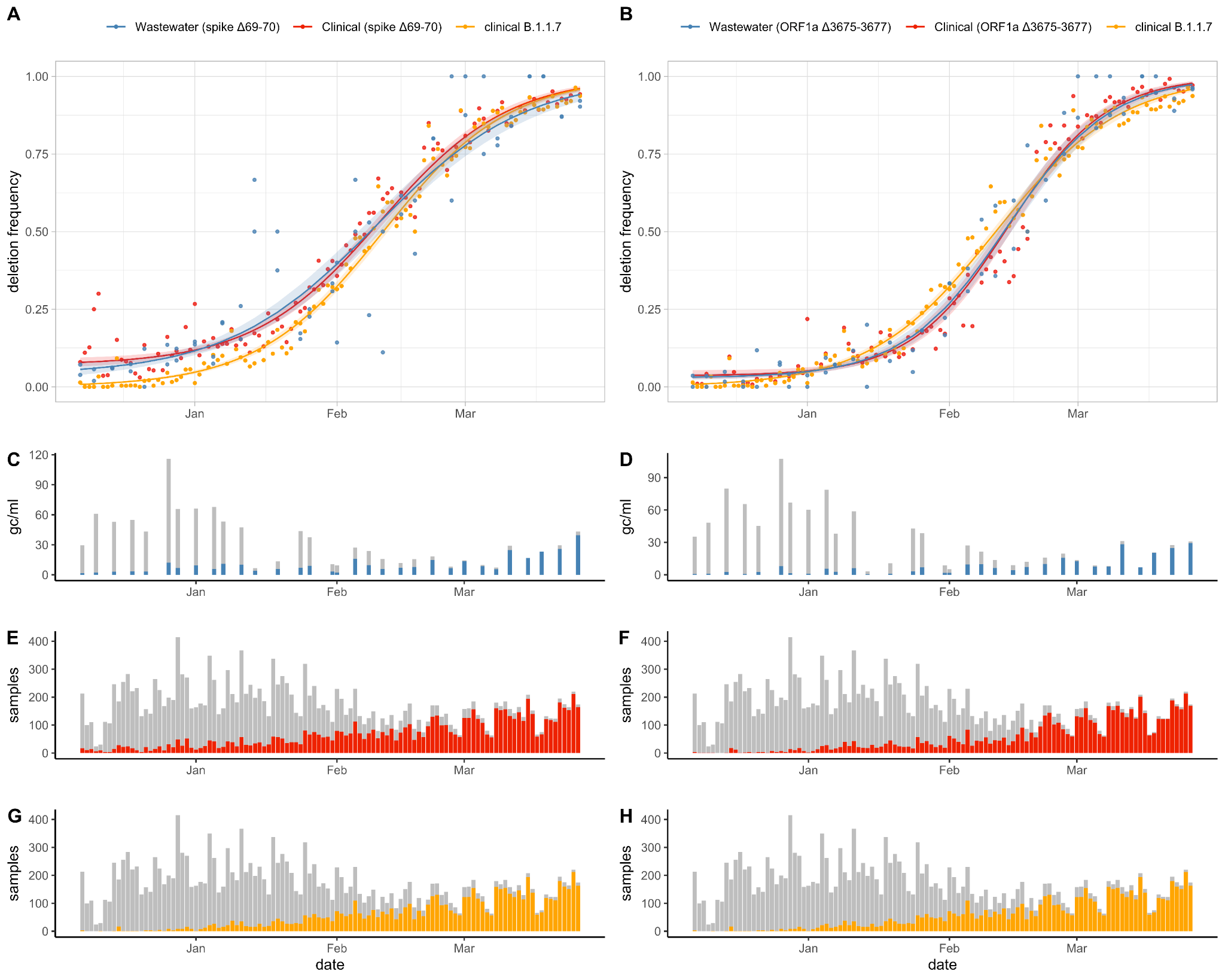

**Figure S4.** Proportion of (A) spike Δ69-70 and (B) ORF1a Δ3675-3677 in wastewater samples compared to clinical samples from throughout Switzerland, along with the proportion of B.1.1.7 lineage in clinical samples from throughout Switzerland. Fitted curves correspond to three-parametric logistic fits for the deletions in wastewater and clinical data and two-parametric logistic fit for the B.1.1.7 clinical data. Shaded areas correspond to 95% confidence bands. Concentration of SARS-CoV-2 RNA (grey) and concentration of deletion alleles (blue) for the (C) spike Δ69-70 and (D) ORF1a Δ3675-3677 in wastewater samples. Number of Switzerland sequenced clinical samples (grey) and number of samples with the deleted allele (red) for the (E) spike Δ69-70 and (F) ORF1a Δ3675-3677. (G, H) Number of Switzerland sequenced clinical samples (grey) and number of samples from the B.1.1.7 lineage (orange).

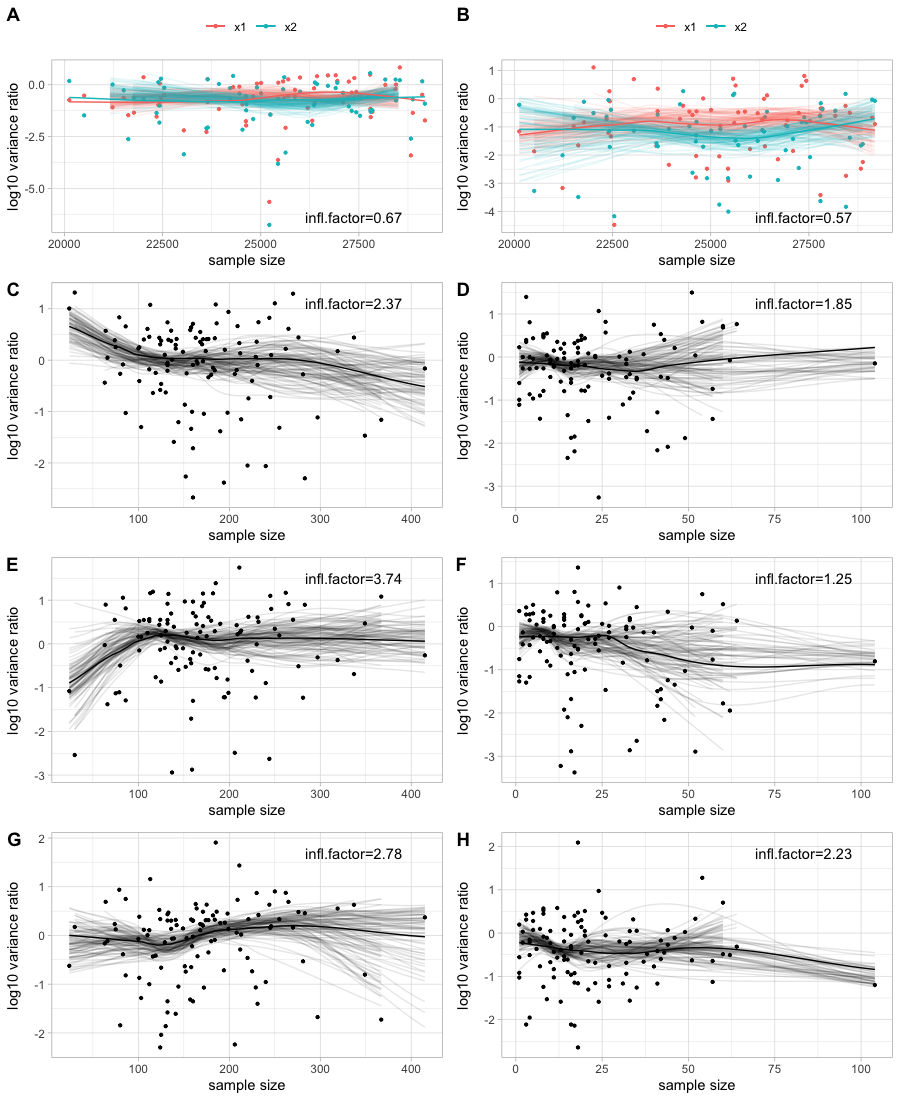

**Figure S5.** Plots of log10 overdispersion vs sample size showing that the assumption of constant overdispersion of quasilikelihood models holds. Lines are loess smoothed values of all points (bold) and of 100 resamples with replacement (thin). (A) 3PL model for the spike Δ69-70 deletion dPCR data (B) 3PL model for the ORF1a Δ3675-3677 dPCR data (C) 3PL model for the spike Δ69-70 deletion Swiss clinical data (D) 3PL model for the spike Δ69-70 deletion Zurich clinical data (E) 3PL model for the ORF1a Δ3675-3677 Swiss clinical data (F) 3PL model for the ORF1a Δ3675-3677 Zurich clinical data (G) 2PL model for the B.1.1.7 Swiss clinical data (H) 2PL model for the B.1.1.7 Zurich clinical data

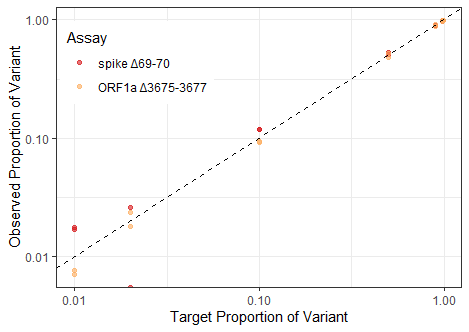

**Figure S6.** The drop-off RT-dPCR assays allow estimation of the proportion of amplicons containing either the spike Δ69-70 (red) or ORF1a Δ3675-3677 (orange) deletions. Target proportion of variants tested included 0.01, 0.02, 0.1, 0.5, 0.98, and 0.99. The dashed line represents expectation with a slope of 1. Axes are log-transformed for visualization.

**Table S2.** Three parametric (3PL) and two-parametric (2PL) logistic model parameter estimates for the prevalence of spike Δ69-70 and ORF1a Δ3675-3677 in wastewater data, Swiss clinical data and Zurich clinical data, as well as for the prevalence of B.1.1.7 variants in Swiss and Zurich clinical data. Values are maximum likelihood estimates and Wald 95% confidence intervals of the growth rate $a$, midpoint $t_{0}$ (in days after December 07, 2020) and (in the case of 3PL) background prevalence $c$. Values for the rate parameter $a$ are also shown transformed (along with their confidence intervals) into an estimate of the transmission fitness advantage $f_{d}$ assuming the discrete-time growth model found in Chen et al. (Chen, Nadeau, Topolsky, Manceau, Huisman, Jablonski, et al. 2021). Last column shows the p-value of a likelihood ratio test (LRT) comparing the 3PL and the 2PL (using the quasibinomial or quasimultinomial quasilikelihoods), assuming a chi-square distribution of the statistic under the null.

| **Sample Type** | ***Growth Rate* (**$a$***)*** | **Time to Maximum Growth *(***$t_{0}$***)*** | **Background Prevalenc*e (***$c$***)*** | **Transmission Fitness Advantage *(***$f_{d}$***)*** | ***LRT p-value*** |
| --- | --- | --- | --- | --- | --- |
| wastewater spike Δ69-70 3PL | 0.06 [0.06, 0.07] | 64.5 [62.0, 67.1] | 0.04 [0.01, 0.06] | 0.34 [0.30, 0.39] | <1e-16 |
| wastewater spike Δ69-70 2PL | 0.05 [0.05, 0.06] | 61.8 [60.0, 63.7] | 0 (fixed) | 0.3 [0.28, 0.32] |  |
| wastewater ORF1a Δ3675-3677 3PL | 0.09 [0.08, 0.09] | 68.5 [67.4, 69.6] | 0.03 [0.02, 0.04] | 0.53 [0.49, 0.57] | 2.08e-06 |
| wastewater ORF1a Δ3675-3677 2PL | 0.07 [0.07, 0.07] | 67.1 [65.6, 68.5] | 0 (fixed) | 0.4 [0.38, 0.43] |  |
| Switzerland spike Δ69-70 3PL | 0.07 [0.06, 0.08] | 65.5 [63.5, 67.6] | 0.07 [0.04, 0.09] | 0.41 [0.36, 0.46] | 5.38e-09 |
| Switzerland spike Δ69-70 2PL | 0.06 [0.05, 0.06] | 61.6 [58.4, 64.8] | 0 (fixed) | 0.32 [0.27, 0.36] |  |
| Switzerland ORF1a Δ3675-3677 3PL | 0.09 [0.08, 0.11] | 68.7 [66.1, 71.2] | 0.03 [0, 0.05] | 0.56 [0.45, 0.67] | 3.77e-05 |
| Switzerland ORF1a Δ3675-3677 2PL | 0.08 [0.07, 0.09] | 66.9 [63.5, 70.2] | 0 (fixed) | 0.46 [0.38, 0.54] |  |
| Switzerland B.1.1.7 3PL | 0.07 [0.07, 0.08] | 66.1 [64.2, 68.0] | 0 [-0.01, 0.01] | 0.42 [0.37, 0.47] | 9.99e-01 |
| Switzerland B.1.1.7 2PL | 0.07 [0.07, 0.08] | 66.1 [64.4, 67.8] | 0 (fixed) | 0.42 [0.38, 0.46] |  |
| Zurich spike Δ69-70 3PL | 0.09 [0.07, 0.11] | 76.3 [71.8, 80.9] | 0.11 [0.07, 0.14] | 0.55 [0.39, 0.73] | 1.77e-07 |
| Zurich spike Δ69-70 2PL | 0.05 [0.03, 0.06] | 72.0 [61.5, 82.5] | 0 (fixed) | 0.24 [0.16, 0.33] |  |
| Zurich ORF1a Δ3675-3677 3PL | 0.09 [0.08, 0.1] | 71.3 [68.8, 73.7] | 0.01 [0, 0.03] | 0.55 [0.46, 0.65] | 1.72e-02 |
| Zurich ORF1a Δ3675-3677 2PL | 0.08 [0.07, 0.09] | 70.7 [67.0, 74.5] | 0 (fixed) | 0.47 [0.39, 0.56] |  |
| Zurich B.1.1.7 3PL | 0.09 [0.07, 0.1] | 74.8 [71.7, 77.9] | 0.01 [-0.01, 0.02] | 0.54 [0.43, 0.65] | 2.01e-01 |
| Zurich B.1.1.7 2PL | 0.08 [0.07, 0.1] | 74.6 [70.0, 79.2] | 0 (fixed) | 0.49 [0.38, 0.61] |  |
